# Association Between Neutrophil-to-Lymphocyte Ratio and Cardiac Sympathetic Denervation in Parkinson’s Disease

**DOI:** 10.64898/2026.10.18.26365787

**Authors:** Makoto Takahashi, Kei Yoshino, Ryota Nozaki, Takashi Yamaguchi, Keita Taguchi, Yoshio Suzuki, Kai Takahashi, Kenta Takahashi, Kazuhiro Iwaoka, Tetsuya Maeda

**Author notes:** Corresponding author: Dr. Makoto Takahashi., Division of Neurology and Gerontology, Department of Internal Medicine, School of Medicine, Iwate Medical University, Iwate, Japan., 2-1-1 Idaidori, Yahaba-cho, Siwa-gun, Iwate 028-3609, japan, (MT).

## Abstract

**Background:** Peripheral inflammation has been implicated in the pathophysiology of Parkinson’s disease (PD), but its relationship with autonomic dysfunction remains unclear.

**Objectives:** To explore the association between peripheral inflammation, assessed by the neutrophil-to-lymphocyte ratio (NLR), and autonomic function in patients with PD.

**Methods:** We retrospectively analyzed 97 patients with PD who underwent ^123^I-metaiodobenzylguanidine (MIBG) cardiac scintigraphy and differential blood cell counts. Associations of NLR with early and delayed heart-to-mediastinum (H/M) ratios, washout ratio, and the coefficient of variation of R-R intervals (CVRR) were assessed using Spearman’s rank correlation and multivariable linear regression adjusted for age, sex, and disease duration.

**Results:** No significant univariable correlations were observed between NLR and the autonomic measures. In multivariable analysis, higher NLR was associated with a lower early H/M ratio (B = −0.802, 95% CI −1.598 to −0.005, P = 0.049), whereas no significant associations were observed with the delayed H/M ratio, washout ratio, or CVRR. Sensitivity analyses showed a generally consistent direction of association, although the effect estimate was attenuated after exclusion of two influential observations and statistical significance was not consistently retained.

**Conclusions:** Higher NLR was associated with lower early cardiac MIBG uptake after adjustment for age, sex, and disease duration in patients with PD. This exploratory finding raises the possibility of an association between systemic inflammation and sympathetic dysfunction in PD and requires confirmation in larger prospective studies.

## Introduction

Parkinson’s disease (PD) is a neurodegenerative disorder characterized not only by motor symptoms but also by a broad range of nonmotor manifestations, including autonomic dysfunction (1). Autonomic dysfunction can occur during the prodromal or early stages of PD, and early autonomic dysfunction has been associated with more rapid disease progression and shorter survival (2).

Accumulating evidence suggests that chronic inflammation and immune dysregulation are involved in the pathophysiology of PD, not only in the central nervous system but also in the periphery (3). The neutrophil-to-lymphocyte ratio (NLR) is a readily available marker of systemic inflammation, and previous studies have reported higher NLR in patients with PD than in healthy controls (4).

Importantly, the autonomic nervous system plays a role in regulating peripheral immune responses through neural–immune interactions. Both sympathetic and parasympathetic pathways are involved in the modulation of inflammatory responses and maintenance of immune homeostasis (5). These observations raise the possibility that autonomic dysfunction and peripheral inflammation may be linked in PD. However, the relationship between peripheral inflammatory markers and autonomic dysfunction in PD remains poorly understood.

Therefore, we explored the association between NLR and autonomic function in patients with PD, using ^123^I-metaiodobenzylguanidine (MIBG) cardiac scintigraphy, an in vivo marker of cardiac sympathetic denervation (6), and the coefficient of variation of R-R intervals (CVRR), a measure of cardiac parasympathetic function (7), as autonomic indices.

## Materials and Methods

### Ethics Statement

The study was approved by the Ethics Committee of Iwate Medical University (approval number: MH2026-018). Because of the retrospective nature of the study, participants were provided with the opportunity to opt out of the study.

### Study Design and Participants

This retrospective observational study included patients with PD who were admitted to our hospital between April 2021 and March 2026, met the Movement Disorder Society Clinical Diagnostic Criteria for Parkinson’s Disease (8), and underwent both ^123^I-MIBG cardiac scintigraphy and differential blood cell counts during the same hospitalization. Patients with conditions or medications that could substantially affect systemic inflammation, autonomic function, or cardiac MIBG uptake were excluded. Exclusion criteria included active infection, malignancy, autoimmune disease, diabetes mellitus of more than 10 years’ duration, current insulin therapy, a history of myocardial infarction or thoracic surgery, regular use of nonsteroidal anti-inflammatory drugs, systemic corticosteroid therapy, and immunosuppressive therapy.

### Assessment of Peripheral Inflammation

The NLR was calculated from differential blood cell counts obtained on admission by dividing the absolute neutrophil count by the absolute lymphocyte count.

### Assessment of Autonomic Function

^123^I-MIBG cardiac scintigraphy was performed during the same hospitalization as blood sampling, generally within several days. After intravenous administration of 128 MBq of ^123^I-MIBG, planar images were obtained at 30 min (early phase) and 3 h (delayed phase). Early and delayed heart-to-mediastinum (H/M) ratios and the washout ratio were calculated using semiautomated software.

CVRR was measured from resting electrocardiograms obtained during hospitalization. One hundred consecutive R-R intervals were recorded on three occasions, and the mean of the valid measurements was used for analysis. Patients with arrhythmias that precluded reliable measurement were excluded from the CVRR analysis.

### Statistical Analysis

Continuous variables are presented as mean ± standard deviation or median with interquartile range (IQR), as appropriate. Associations between NLR and autonomic function measures, including early and delayed H/M ratios, washout ratio, and CVRR, were initially assessed using Spearman’s rank correlation coefficients. Multivariable linear regression analyses were subsequently performed with NLR as the dependent variable and each autonomic function measure as an independent variable, adjusting for age, sex, and disease duration. Sensitivity analyses for the association between NLR and the early H/M ratio were performed by additionally adjusting for Hoehn and Yahr stage, log-transforming NLR, excluding influential observations identified using Cook’s distance (>4/n), and applying HC3 robust standard errors. Given the exploratory nature of the study, no adjustment for multiple comparisons was performed. A two-sided P value <0.05 was considered statistically significant. All statistical analyses were performed using EZR (9).

## Results

### Patient Characteristics

During the study period, 109 consecutive patients met the inclusion criteria. Twelve patients met the exclusion criteria, leaving 97 patients for the final analysis (Table 1). Each patient was included only once. The mean age was 65.4 ± 10.2 years, and 42 patients (43.3%) were male. The median disease duration was 3.0 years (IQR, 2.0–10.0 years). The numbers of patients with Hoehn and Yahr stages 2, 3, 4, and 5 were 26, 52, 17, and 2, respectively. The mean NLR was 2.74 ± 1.23. The mean early and delayed H/M ratios were 1.80 ± 0.34 and 1.60 ± 0.42, respectively, and the mean washout ratio was 47.5 ± 17.1%. CVRR was available in 70 patients, as it was not performed in 24 patients and three patients were excluded because of arrhythmias that precluded reliable measurement, with a mean value of 1.99 ± 1.05%.

**Table 1.** Clinical characteristics of the study participants.

| table 1 |  |
| --- | --- |
| (N=97) | mean |
| Age, years | $65.4 \pm 10.2$ |
| Disease duration, years | 3.0 [2.0-10.0] |
| sex, male/female, n (%) | 42 (43.3) / 55 (56.7) |
| HY2, n (%) | 26 (26.8) |
| HY3, n (%) | 52 (53.6) |
| HY4, n (%) | 17 (17.5) |
| HY5, n (%) | 2 (2.1) |
| WBC, /ul | $5549.4 \pm 1515.7$ |
| Neutrophil, /ul | $3617.5 \pm 1182.3$ |
| Lymphocyte, /ul | $1462.9 \pm 551.6$ |
| NLR | $2.74 \pm 1.23$ |
| H/M early | $1.802 \pm 0.339$ |
| H/M delay | $1.602 \pm 0.424$ |
| WR | $47.5 \pm 17.1$ |
| CVRR mean, % (n=70) | $1.99 \pm 1.05$ |
| Table 1. Clinical characteristics of the study participants |  |
| Data are presented as mean ± standard deviation or median [interquartile range], unless otherwise indicated. | |
| Abbreviations: CVRR, coefficient of variation of R-R intervals; H/M, heart-to-mediastinum; HY, Hoehn and Yahr stage; NLR, neutrophil-to-lymphocyte ratio; WBC, white blood cell count; WR, washout ratio. |  |

### Univariate Analysis

Spearman’s rank correlation analysis showed no significant correlations between NLR and the early H/M ratio (ρ = −0.147, P = 0.152), delayed H/M ratio (ρ = −0.101, P = 0.324), washout ratio (ρ = 0.063, P = 0.539), or CVRR (ρ = −0.100, P = 0.410) (Figure 1).

**Figure 1.**
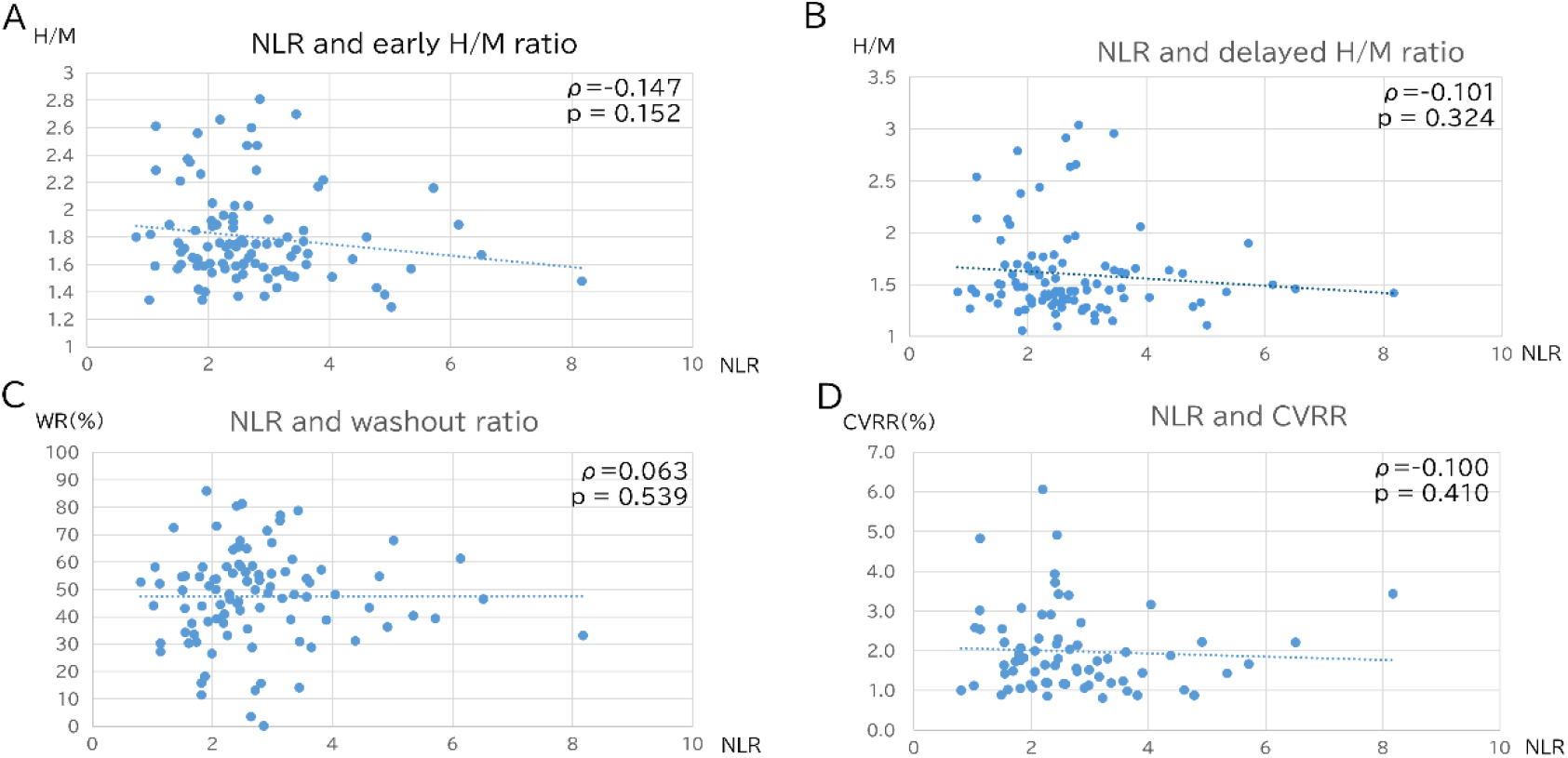
Associations between the neutrophil-to-lymphocyte ratio (NLR) and autonomic function measures in patients with Parkinson’s disease. Scatter plots show the associations of the NLR with (A) early heart-to-mediastinum (H/M) ratio, (B) delayed H/M ratio, (C) MIBG washout ratio, and (D) coefficient of variation of R-R intervals (CVRR). Spearman’s rank correlation coefficients (ρ) and P values are shown in each panel.

### Multivariable Analysis

In multivariable linear regression analyses adjusted for age, sex, and disease duration, NLR was inversely associated with the early H/M ratio (B = −0.802, 95% CI −1.598 to −0.005, P = 0.049) (Table 2). No significant associations were observed with the delayed H/M ratio, washout ratio, or CVRR. In sensitivity analyses, the effect estimate for the early H/M ratio remained largely unchanged after additional adjustment for Hoehn and Yahr stage and when HC3 robust standard errors were applied. However, exclusion of two influential observations attenuated the regression coefficient from −0.802 to −0.557, and statistical significance was not retained (Table 3).

**Table 2.** Multivariable linear regression analyses of the association between NLR and autonomic function measures.

| Explanatory variables | B | 95% CI | p |
| --- | --- | --- | --- |
| H/M early (n=97) | -0.802 | -1.598 ~ -0.005 | 0.049 |
| H/M delay (n=97) | -0.45 | -1.087 ~ 0.188 | 0.164 |
| WR (n=97) | 0.002 | -0.013 ~ 0.018 | 0.769 |
| CVRR mean (n=70) | -0.032 | -0.406 ~ 0.342 | 0.864 |

NLR was the dependent variable. Each autonomic function measure was entered separately as an independent variable, with adjustment for age, sex, and disease duration.
Abbreviations: B, unstandardized regression coefficient; CI, confidence interval; CVRR, coefficient of variation of R-R intervals; H/M, heart-to-mediastinum; NLR, neutrophil-to-lymphocyte ratio; WR, washout ratio.

**Table 3.**
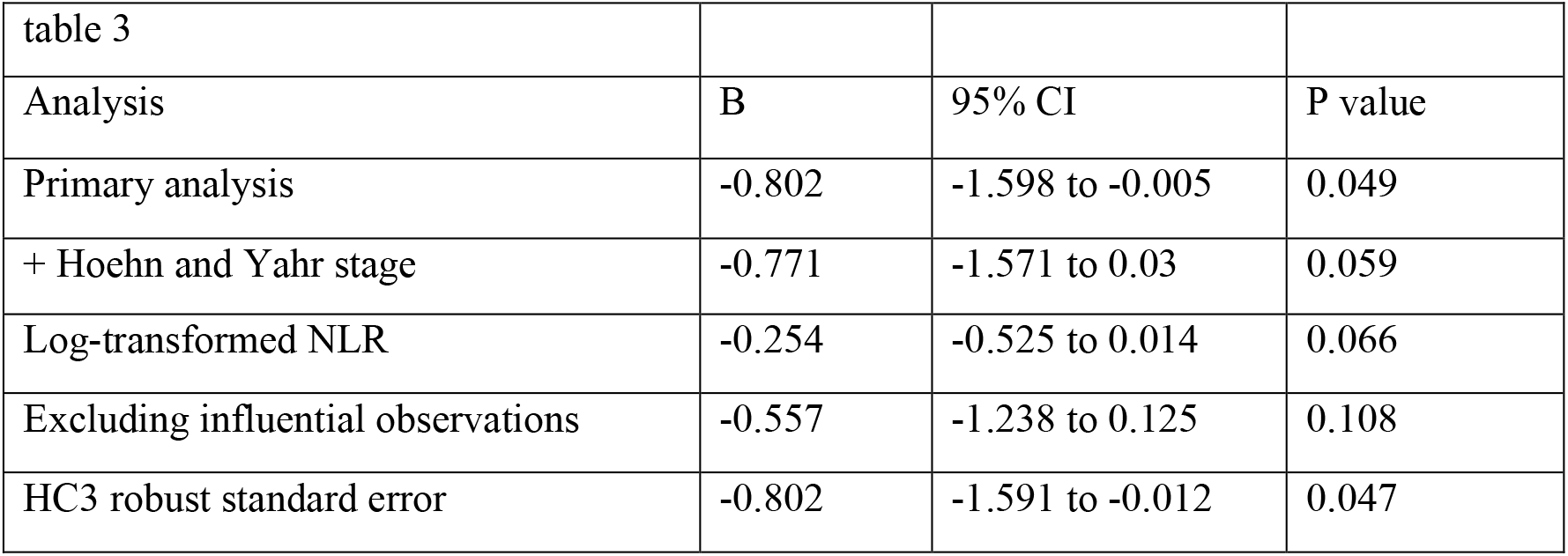
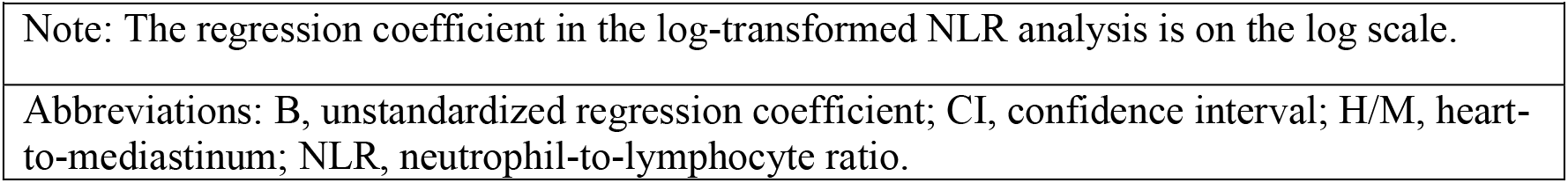
Sensitivity analyses of the association between NLR and the early H/M ratio.

| Analysis | B | 95% CI | P value |
| --- | --- | --- | --- |
| Primary analysis | -0.802 | -1.598 to -0.005 | 0.049 |
| + Hoehn and Yahr stage | -0.771 | -1.571 to 0.03 | 0.059 |
| Log-transformed NLR | -0.254 | -0.525 to 0.014 | 0.066 |
| Excluding influential observations | -0.557 | -1.238 to 0.125 | 0.108 |
| HC3 robust standard error | -0.802 | -1.591 to -0.012 | 0.047 |

|  |
| --- |
| Table 3. Sensitivity analyses of the association between NLR and the early H/M ratio |
| Note: The regression coefficient in the log-transformed NLR analysis is on the log scale. |
| Abbreviations: B, unstandardized regression coefficient; CI, confidence interval; H/M, heart-to-mediastinum; NLR, neutrophil-to-lymphocyte ratio. |

## Discussion

In this retrospective study of patients with PD, NLR was not significantly correlated with any of the autonomic measures in univariable analyses. After adjustment for age, sex, and disease duration, however, a higher NLR was associated with a lower early H/M ratio on ^123^I-MIBG cardiac scintigraphy, whereas no significant associations were observed with the delayed H/M ratio, washout ratio, or CVRR. These findings suggest a possible link between systemic inflammation and sympathetic dysfunction in PD, although the observed association was exploratory and should be interpreted cautiously.

The sympathetic nervous system represents a potential link between peripheral inflammation and autonomic dysfunction, as catecholaminergic signaling is involved in the neural regulation of immune responses (5). Sympathetic noradrenergic dysfunction is a well-established feature of PD, with cardiac sympathetic denervation being particularly prominent (6,8). Therefore, the inverse association observed between NLR and the early H/M ratio may reflect an interaction between peripheral inflammatory activity and sympathetic dysfunction in PD. However, the present cross-sectional findings cannot determine the direction or mechanism of this relationship.

The association with NLR was observed for the early H/M ratio but not for the delayed H/M ratio. We previously demonstrated that both early and delayed cardiac MIBG uptake correlated quantitatively with the residual cardiac sympathetic axons in autopsy-confirmed Lewy body disease (6), suggesting that the two measures should not necessarily be interpreted as reflecting distinct aspects of sympathetic dysfunction. Therefore, the discrepancy between the early and delayed findings in the present study should not necessarily be interpreted as reflecting distinct pathophysiological processes. The reason for this discrepancy remains unclear and requires further investigation.

No significant associations were observed between NLR and either the MIBG washout ratio or CVRR. CVRR has been used as a measure of cardiac parasympathetic function in patients with PD (7). The differing findings for cardiac MIBG uptake and CVRR may reflect differences in the autonomic components assessed by these measures. However, the absence of an association with CVRR should not be interpreted as evidence of a specific relationship between NLR and sympathetic dysfunction, particularly because CVRR was available in only 70 patients.

It should also be noted that the association between NLR and the early H/M ratio emerged only after adjustment for age, sex, and disease duration and was not evident in the univariable analysis. Sensitivity analyses showed generally stable effect estimates, although exclusion of two influential observations attenuated the regression coefficient from −0.802 to −0.557 (Table S1), suggesting that the estimated magnitude of the association may have been influenced by a small number of patients. Furthermore, because multiple autonomic measures were examined without adjustment for multiple comparisons, the association with the early H/M ratio may represent a chance finding. Accordingly, the present finding should be regarded as exploratory and requires confirmation in an independent cohort.

Several limitations should be acknowledged. First, this was a retrospective, cross-sectional, single-center study with a relatively small sample size, precluding assessment of causal or temporal relationships. In addition, the diagnosis of PD was based on clinical criteria without pathological confirmation. Second, NLR is a nonspecific marker of systemic inflammation and may be influenced by unmeasured factors despite the exclusion of major potential confounders. Third, cardiac MIBG scintigraphy reflects cardiac sympathetic denervation and may not represent sympathetic involvement in other organs. In addition, CVRR was available in only a subset of patients, which may have reduced the power to detect an association. Prospective studies incorporating more specific inflammatory markers and multiple measures of autonomic function are needed to confirm these findings and clarify their pathophysiological significance.

In conclusion, higher NLR was associated with lower early cardiac MIBG uptake after adjustment for age, sex, and disease duration in patients with PD. Although exploratory, these findings raise the possibility of an association between systemic inflammation and cardiac sympathetic denervation in PD and warrant confirmation in larger prospective studies.

## Data Availability

The datasets analyzed during the current study are not publicly available due to patient privacy considerations but may be available from the corresponding author upon reasonable request, subject to institutional and ethical approval.

## Acknowledgments

The authors would like to thank Ms. Akie Sakamoto and Ms. Kunie Watanabe for their assistance with sample record management.

## Author Contribution

Conceptualization: MT

Formal analysis: MT

Investigation: KY, RN, TY, KeiT, YS, KaiT, KenT, and KI

Writing-original draft: MT

Writing-review & editing: MT and TM

## Availability of Data

The data that support the findings of this study are available from the corresponding author upon reasonable request.

Declaration of generative AI and AI-assisted technologies in the manuscript preparation process During the preparation of this work, the authors used ChatGPT (OpenAI) to assist with language editing and improving the clarity and organization of the manuscript. After using this tool, the authors reviewed and edited the content as needed and take full responsibility for the content of the published article.

## References

1. Kalia LV, Lang AE. Parkinson’s disease. The Lancet. 2015 Aug;386(9996):896–912. doi:10.1016/S0140-6736(14)61393-3

2. De Pablo-Fernandez E, Tur C, Revesz T, Lees AJ, Holton JL, Warner TT. Association of Autonomic Dysfunction With Disease Progression and Survival in Parkinson Disease. JAMA Neurol. 2017 Aug 1;74(8):970. doi:10.1001/jamaneurol.2017.1125

3. Lauritsen J, Romero-Ramos M. The systemic immune response in Parkinson’s disease: focus on the peripheral immune component. Trends Neurosci. 2023 Oct;46(10):863–78. doi:10.1016/j.tins.2023.07.005

4. Cabanillas-Lazo M, Montes-Baldarrago A, Uriol-Alvino S, Baltazar-Ñahui H, Pereda-Leon NB, Jimenez-Alvarez H, et al. The neutrophil-to-lymphocyte ratio in Parkinson’s disease: a systematic review and meta-analysis of its association with differential diagnosis, severity, and prognosis. Clin Park Relat Disord. 2026;15:100494. doi:10.1016/j.prdoa.2026.100494

5. Pavlov VA, Tracey KJ. Neural regulation of immunity: molecular mechanisms and clinical translation. Nat Neurosci. 2017 Feb;20(2):156–66. doi:10.1038/nn.4477

6. Takahashi M, Ikemura M, Oka T, Uchihara T, Wakabayashi K, Kakita A, et al. Quantitative correlation between cardiac MIBG uptake and remaining axons in the cardiac sympathetic nerve in Lewy body disease. J Neurol Neurosurg Psychiatry. 2015 Sep;86(9):939–44. doi:10.1136/jnnp-2015-310686

7. Miyagi T, Yamazato M, Nakamura T, Tokashiki T, Namihira Y, Kokuba K, et al. Power spectral analysis of heart rate variability is useful as a screening tool for detecting sympathetic and parasympathetic nervous dysfunctions in Parkinson’s disease. BMC Neurol. 2022 Sep 10;22(1):339. doi:10.1186/s12883-022-02872-2

8. Postuma RB, Berg D, Stern M, Poewe W, Olanow CW, Oertel W, et al. MDS clinical diagnostic criteria for Parkinson’s disease. Mov Disord. 2015 Oct;30(12):1591–601. doi:10.1002/mds.26424

9. Kanda Y. Investigation of the freely available easy-to-use software ‘EZR’ for medical statistics. Bone Marrow Transplant. 2013 Mar;48(3):452–8. doi:10.1038/bmt.2012.244

